# Low frequency interictal EEG biomarker for localizing seizures

**DOI:** 10.1101/2021.06.04.21258382

**Authors:** Brian Nils Lundstrom, Benjamin Brinkmann, Gregory Worrell

## Abstract

**Objective:** We want to identify seizure onset zone (SOZ) from interictal EEG biomarkers. We hypothesize that a combination of interictal EEG biomarkers, including a novel low frequency marker, can predict mesial temporal involvement and can assist in prognosis related to surgical resections.

**Methods:** Interictal direct current wide bandwidth invasive EEG recordings from 83 patients implanted with 5,111 electrodes were retrospectively studied. Logistic regression was used to classify electrodes and patient outcomes. A feed-forward neural network was implemented to understand putative mechanisms.

**Results:** Interictal infraslow frequency EEG activity was decreased for SOZ electrodes while faster frequencies such as delta (2-4 Hz) and beta-gamma (20-50 Hz) activity were increased. These spectral changes comprised a novel interictal EEG biomarker that was significantly increased for mesial temporal SOZ electrodes compared to non-SOZ electrodes. Interictal EEG biomarkers correctly classified mesial temporal SOZ electrodes with a specificity of 87% and positive predictive value of 80%. These interictal EEG biomarkers also correctly classified patient outcomes after surgical resection with a specificity of 91% and positive predictive value of 87%.

**Interpretation:** Interictal infraslow EEG activity is decreased near the SOZ while higher frequency power is increased, suggesting distinct underlying physiologic mechanisms. Decreased interictal infraslow activity may reflect the loss of neural inhibition. Narrowband interictal EEG power bands provide information about the SOZ and can help predict mesial temporal involvement in seizure onset. Together with interictal epileptiform discharges and high frequency oscillations, these interictal biomarkers may provide prognostic information prior to surgical resection.

## Introduction

Approximately one-third of epilepsy patients are resistant to anti-seizure medications and are candidates for second-line treatments including surgical resection or neuromodulation. Often it is imperative to precisely localize the epileptogenic zone, the region of brain tissue that generates spontaneous unprovoked seizures. This is typically accomplished by determining the location from which seizures arise, or the seizure onset zone (SOZ), with intracranial electroencephalographic (EEG) monitoring. Appropriately sampling seizures in order to localize the SOZ remains challenging due to the unpredictability of seizures, sparse spatial sampling of EEG, and associated morbidities. In addition, the optimal EEG spectral range for mapping the SOZ remains unclear ^1^.

Improved interictal biomarkers to assist in the localization of the SOZ could decrease patient morbidity and improve outcomes. Prior work on interictal localization of SOZ has focused on interictal epileptiform spikes ^2–4^ and high frequency oscillations (HFOs) ^5–9^. Both spikes and HFOs have been associated with hyperexcitable tissue. In contrast to spikes and HFOs, here we focus on lower frequency activity. The appearance of slow-wave EEG shifts are well-described at the onset of focal seizures ^10^. However, the role of pathological focal interictal activity in epileptogenic brain has received less attention ^11^. Recent work from eight patients suggested that for SOZ electrodes infraslow activity is decreased relative to fast delta activity and gamma (30-50 Hz) activity is increased ^12^.

Single neuron physiology and modeling suggesting that activity less than approximately 4 Hz may be related to neural mechanisms controlling excitability ^13–16^. Low frequency EEG activity has been shown to modulate cortical excitability: Low frequency activity modulates interictal spiking and sleep architecture ^1^, is related to periods of increased and decreased neuronal activity during sleep ^17^, and may be transiently decreased during activation tasks as gamma activity increases ^18^. Delta activity during sleep is correlated with local synaptic strength ^19,20^, and during wakefulness increased focal delta is often considered a marker of cortical dysfunction ^21^. Our hypothesis is that interictal EEG slowing can help localize the seizure onset zone in patients with focal epilepsy.

We examine the utility of these interictal biomarkers to localize seizure onset without ictal data. For example, although unilateral mesial temporal lobe epilepsy can be effectively treated with surgical resection ^22^, a common challenge is to accurately determine whether patients have only unilateral temporal onset seizures. The average time to record the first contralateral seizure in patients with bilateral seizures was approximately six weeks ^23^, longer than typical admissions for invasive monitoring. Patients with bilateral onset seizures can be effectively treated with electrical stimulation ^24^. Here, we find that mesial temporal electrodes can be classified as SOZ electrodes with a high specificity and positive predictive value. Finally, we show that by using the available electrodes for each patient, patient outcome following resective epilepsy surgery can be predicted with high specificity and positive predictive value. Results from a feedforward neural network suggest that infraslow activity is related to in inhibitory neuronal and synaptic mechanisms.

## Methods

### Data Acquisition and Processing

This retrospective analysis was approved by the Mayo Clinic Institutional Review Board. As described previously ^25^, wide-bandwidth intracranial EEG were acquired on a DC-capable (0-5 kHz) Neuralynx (Bozeman, MT) electrophysiology system from standard clinical electrodes during evaluation of patients for epilepsy surgery between October 2005 and February 2014. All patients with DC-coupled invasive EEG recordings (83 patients and 5,111 total electrode contacts) were included. 39 patients had contacts in the mesial temporal brain regions. Depth electrode platinum-iridium contacts (*n* = 485) were 2.3 mm long, 1 mm diameter, and spaced 5 or 10-mm center-to-center. Subdural grid and strip contacts (*n* = 4,626) were 4.0 mm diameter (2.3 mm exposed) with 10 mm center-to-center distance. Referential data (inverted subgaleal electrode as reference) were chosen from between 1 am and 3 am from the first night following electrode implantation. Data were sampled at 32 kHz, filtered using a Bartlett-Hanning window finite impulse response low-pass filter with 1 kHz cutoff, and down-sampled to 5 kHz.

### Electrode Localization

Three-dimensional coordinates were obtained for all electrode contacts in standard space, as previously described ^25^. Briefly, using the Freesurfer image analysis suite (http://surfer.nmr.mgh.harvard.edu/) preoperative MRIs were mapped from patient space to the FreeSurfer average pial surface and co-registered with post-operative CT scans. Electrode locations were manually labeled in BioImage Suite 3 ^26^. Using iELVis ^27^, electrode locations were determined in average FreeSurfer brain space with segmentation labels from the Desikan-Killiany brain atlas ^28^. Electrode segmentation labels were categorized as follows: mesial temporal lobe included labels for amygdala, hippocampus, entorhinal, and para-hippocampal brain regions; non-mesial temporal lobe regions were termed neocortex and included all other gray matter segmented brain regions.

### Electrophysiology analysis

1.5-2-hour epochs of DC-coupled referential EEG data with inverted subgaleal reference were analyzed per patient for EEG power, interictal discharges, and high-frequency oscillations. Candidate seizure onset zone (SOZ) electrodes were determined from the clinical invasive EEG report; SOZ electrodes were determined by a board certified epileptologist (GW) as the earliest EEG change in a clear electrographic seizure. Non-SOZ electrodes were contacts from which no initial seizure activity was observed. For each patient, baseline seizure frequency prior to admission was determined from retrospective chart review. HFOs were identified as previously reported ^25^ using an automated detector ^29^. For analysis of low frequency activity, data were decimated to 250 Hz, and a multi-taper spectrum was computed using Chronux (http://chronux.org/) with time-bandwidth product of 5 and 9 tapers ^30^. Frequency spectra were filtered with a 0.012 Hz median filter and down sampled by 25-50 to facilitate data analysis. Similar to previous work ^25^, interictal spike times and amplitudes were found using an automated detector with standard parameters ^31^, and HFOs were identified using a high specificity and sensitivity automated detector ^29^.

### Classification

We performed classification for two cases: electrodes and patient outcomes. To assess the ability of biomarkers to classify electrodes, biomarker values were used in a logistic regression model (linear terms only) to generate probability scores for each contact and compared to electrode labels, i.e. SOZ or non-SOZ. Features for the model included a Novel Marker (median powerband ratio 2-50 Hz/0.02-0.5 Hz), derived from physiological results. Additional features included the rate of interictal discharges per hour, the rate of high frequency oscillations (HFOs), and the patient’s baseline seizure frequency. Multicollinearity was evaluated with the variance inflation factor ^32^, with values <1.6 suggesting predictor variables are not closely related to one another. The scores and labels were used to create a Receiver Operating Characteristics (ROC) curve. The Area Under the Curve (AUC) is a performance measure of the classifier that reflects trade-offs between sensitivity and specificity. For a given threshold value, particular values for sensitivity, specificity, positive predictive value and negative predictive value are generated. Electrodes were grouped by patient. To classify electrodes for a given patient, model training did not include electrodes for that patient, i.e. leave-one-patient-out cross validation was performed.

To classify patient outcomes, patients were divided into those with good outcomes (seizure-free or only auras, ILAE outcome scale 1 or 2) and poor outcomes (continued seizures, ILAE outcome scale 3 or greater). To obtain a single feature value for each patient, the standard deviation of all electrode values for a given biomarker was used. In all, three features were used: the Novel Marker, interictal epileptiform discharge rates, and the patient’s baseline seizure frequency. Logistic regression then classified patient outcome, which was compared to known outcomes. 10-fold cross-validation was performed across patients.

### Statistical Analysis

Statistical testing was conducted using the non-parametric Wilcoxon rank sum test for equal medians, with distributions considered to be significantly different when *p* < 0.05. Boxplots represent the median value, interquartile range (IQR), and 1.5 x IQR with the horizontal line, rectangle, and whiskers, respectively. Statistical significance for AUC values is reported via the *p*-value for the Chi-squared test between the logistic regression model and a constant model. 95% confidence intervals (bias corrected and accelerated) were obtained by bootstrapping (1,000 to 10,000 iterations), which is a widely accepted statistical resampling technique that relies on random sampling with replacement of the distribution in question to provide a non-parametric and robust confidence interval estimate ^33^.

### Neural Network Modeling

Similar to previously ^15^, the feed-forward neural network consisted of conductance-based Hodgkin-Huxley (HH) neurons and synapses that incorporated facilitation and depression ^34^. Three slow adaptation currents were added to the HH neurons as previously ^13^ with time constants of τ = 0.3, 1, and 6 s and conductances of g_1_= 0.05, g_2_=0.006, and g_3_=0.004 times the leak conductance, respectively. For synapses, parameters were similar to those fitted to physiologic data ^34^ and those used previously ^15^. Facilitation and depression variables (F, d, and D) in the synapses relaxed exponentially to one with time constants of 0.1, 0.7, and 9 s for facilitation, fast depression, and slow depression, respectively. For facilitation, each input spike increased F by 0.2, and for depression each input spike decreased d and D to new values of 0.4d and 0.975D, respectively. The synaptic conductance amplitude (F d D) for each neuron was then summed and filtered by an exponential with a time constant of 300 ms. Equations were solved numerically using fourth-order Runge-Kutta integration with a fixed time step of 0.05 ms. Spike times were identified as the upward crossing of the voltage trace at -10 mV (resting potential = -65 mV). Input to each neuron of the neural network was zero-mean exponential filtered (τ = 1 ms) Gaussian white-noise stimuli with standard deviations of 10 and 16 μA cm^-2^. For the loss of adaption conditions, D=0, g_2_=0, and g_3_=0.

### Data Availability

De-identified data are available upon request.

## Results

Overnight DC-EEG recordings from between 1 am and 3 am for 83 patients (53 women) were included in this retrospective IRB-approved analysis. Forty-five (76%) patients were right-handed, and median age was 40 years (range 5-75 years). Median age of seizure onset was 13 years (range 0-55 years). Prior to implantation the median seizure frequency was 6 seizures per month (range 0.4-360 seizure per month). Of the 83 patients, 61 had post-implant imaging sufficient for electrode localization (Figure 1). 39 patients had electrodes located in the mesial temporal head region, and 56 had electrodes in identifiable neocortex. 48 patients (58%) underwent surgical resection. Median follow-up time was 5.4 years (range 0.3-14.5 years), and 31 (65%) patients were free of disabling seizures with either an ILAE Class 1 or Class 2 response.

**Figure 1:**
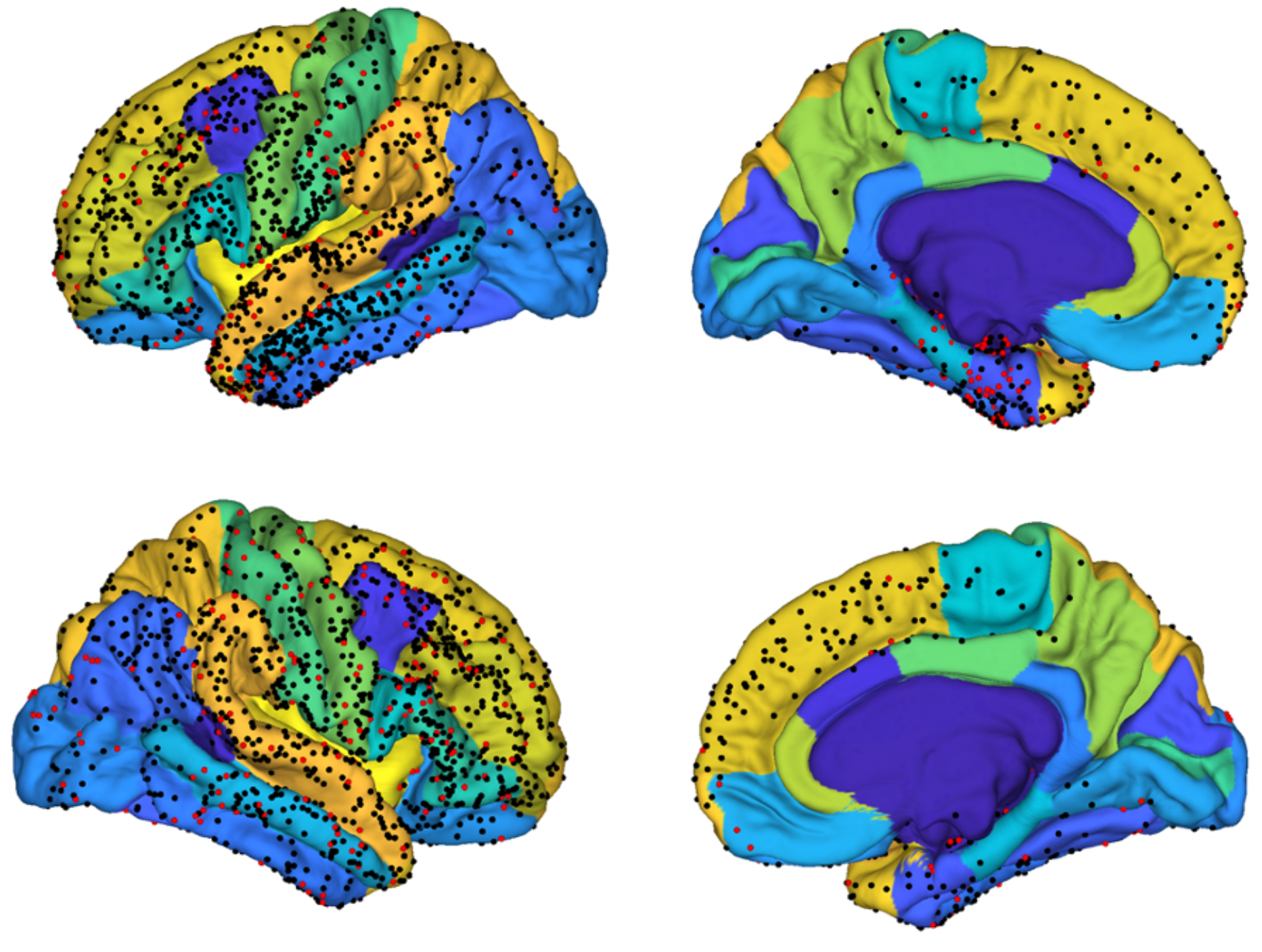
Electrode contact locations from 61 patients on the Desikan-Killiany (D-K) atlas. Seizure onset zone (SOZ) contacts (*n* = 512, red dots) and non-SOZ contacts (*n* = 2,758, black dots) are displayed on the pial surface of the D-K atlas.

Given our prior results from eight patients ^12^, we examined the power spectra of all 5,111 electrodes from 83 patients and found that infraslow activity was decreased for SOZ electrodes compared to non-SOZ electrodes (Figure 2). We used the same powerbands 2-4 Hz and 20-50 Hz. However, here we choose the powerband 0.02-0.5 Hz as 0.02 was the lower limit used in prior work ^1^ and 0.5 Hz is often considered to be the upper limit of infraslow activity (or lower limit of delta frequency activity); we take advantage of DC-coupled amplifiers that were not used in our prior results ^12^. We found that the ratios of powerbands 2-4 Hz to 0.02-0.5 Hz and 20-50 Hz to 0.02-0.5 Hz were significantly increased for SOZ compared to non-SOZ electrodes (Figure 2c).

**Figure 2:**
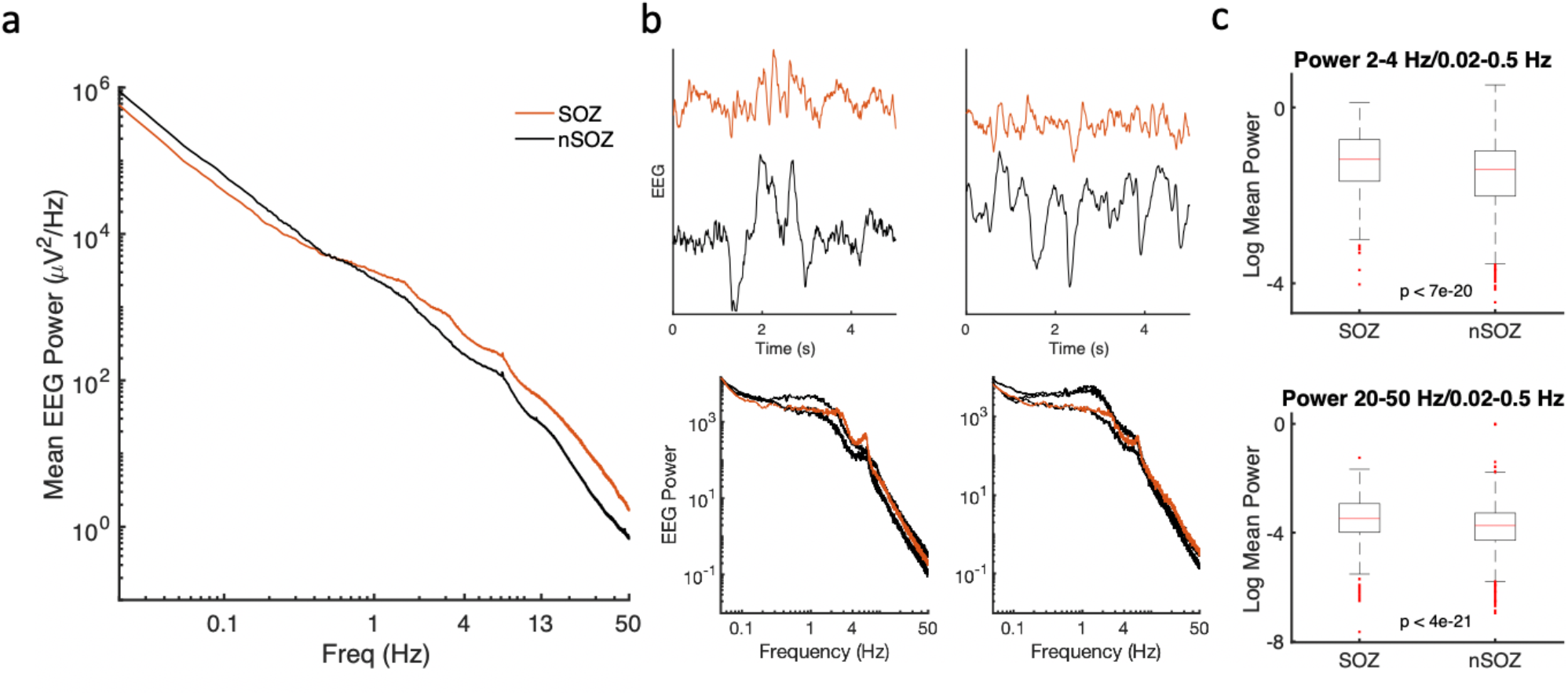
SOZ electrodes show broadband increased EEG power above 1 Hz and decreased lower frequency power. (a) Mean EEG power is increased across a broad range of frequencies for SOZ (*n* = 780) compared to nSOZ electrodes (*n* = 4331) from 83 patients. (b) EEG examples showing SOZ (red) and nearby nSOZ (black) electrodes. (c) For SOZ electrodes, a ratio of fast delta activity (2-4 Hz) to infraslow activity (0.02-0.5 Hz) is increased as well as a ratio of beta-gamma activity (20-50 Hz) to infraslow activity (0.02-0.5 Hz). For smoothing in panel (a), the outlying 1 percentile of powers were removed prior to averaging. Boxplots show the median value (horizontal line), interquartile range (IQR) (rectangle), and 1.5 x IQR (whiskers).

Then, we focused on depth electrodes in the mesial temporal head region and found similar results with decreased infraslow activity and increased beta-gamma activity (Figure 3). With increasing distance from SOZ electrodes, infraslow power increased and the ratio of the of 2-4 Hz and 20-50 Hz power to infraslow power decreased (Figure 3c).

**Figure 3:**
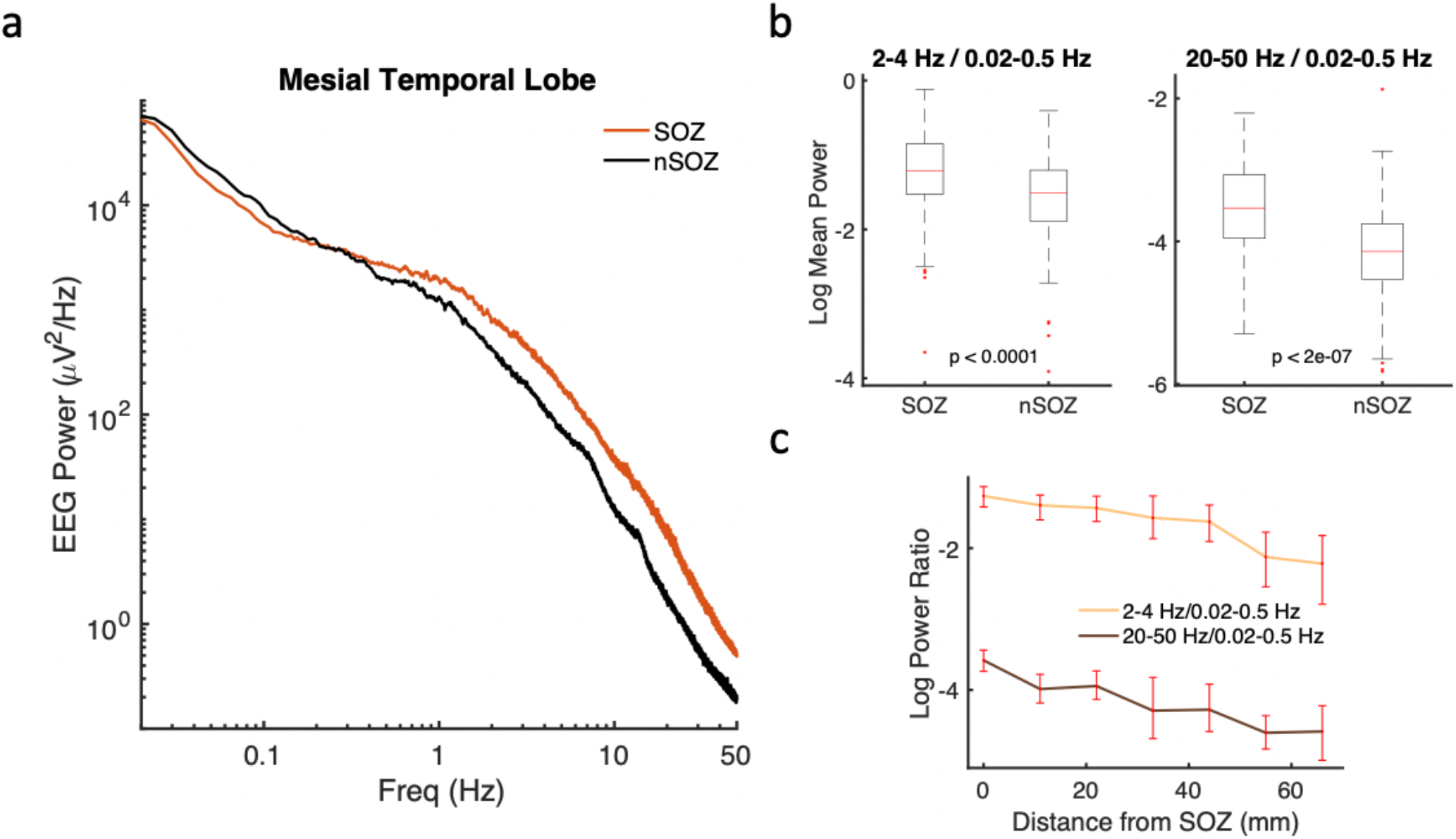
Increased relative fast delta and beta-gamma power from SOZ electrodes is observed in mesial temporal brain regions. (a) Mean EEG power is increased across a broad range of frequencies for SOZ (*n* = 77) compared to nSOZ electrodes (*n* = 85) while infraslow activity is decreased. (b) SOZ electrodes show increased fast delta (2-4 Hz) and increased beta-gamma (20-50 Hz) activity compared to infraslow (0.02-0.5 Hz) activity. (c) These ratios decrease with distance away from the SOZ. Error bars are bootstrapped 95% confidence intervals.

Next, we wanted to see whether these power differences between SOZ and non-SOZ electrodes could help classify electrodes. We constructed a straightforward logistic regression classifier based on interictal EEG features: a Novel Marker (powerband ratio 2-4 Hz/20-50 Hz), interictal epileptiform discharges, and high frequency oscillations. Using all mesial temporal electrodes (*n* = 163) (Figure 4a), results of the classifier are shown for the three features separately as well as with all three features together (Figure 4b). Spikes, HFOs, and the Novel Marker all perform similarly alone but show improved results when combined, which suggests that each contributes distinct information to the classifier; higher AUC values reflect improved overall performance (*p<*0.001 for each value). There is a tradeoff between sensitivity and specificity such that as the decision threshold increases specificity increases and sensitivity decreases (Figure 4c); a threshold of 0.5 yielded a sensitivity of 72% and specificity of 86%.

**Figure 4:**
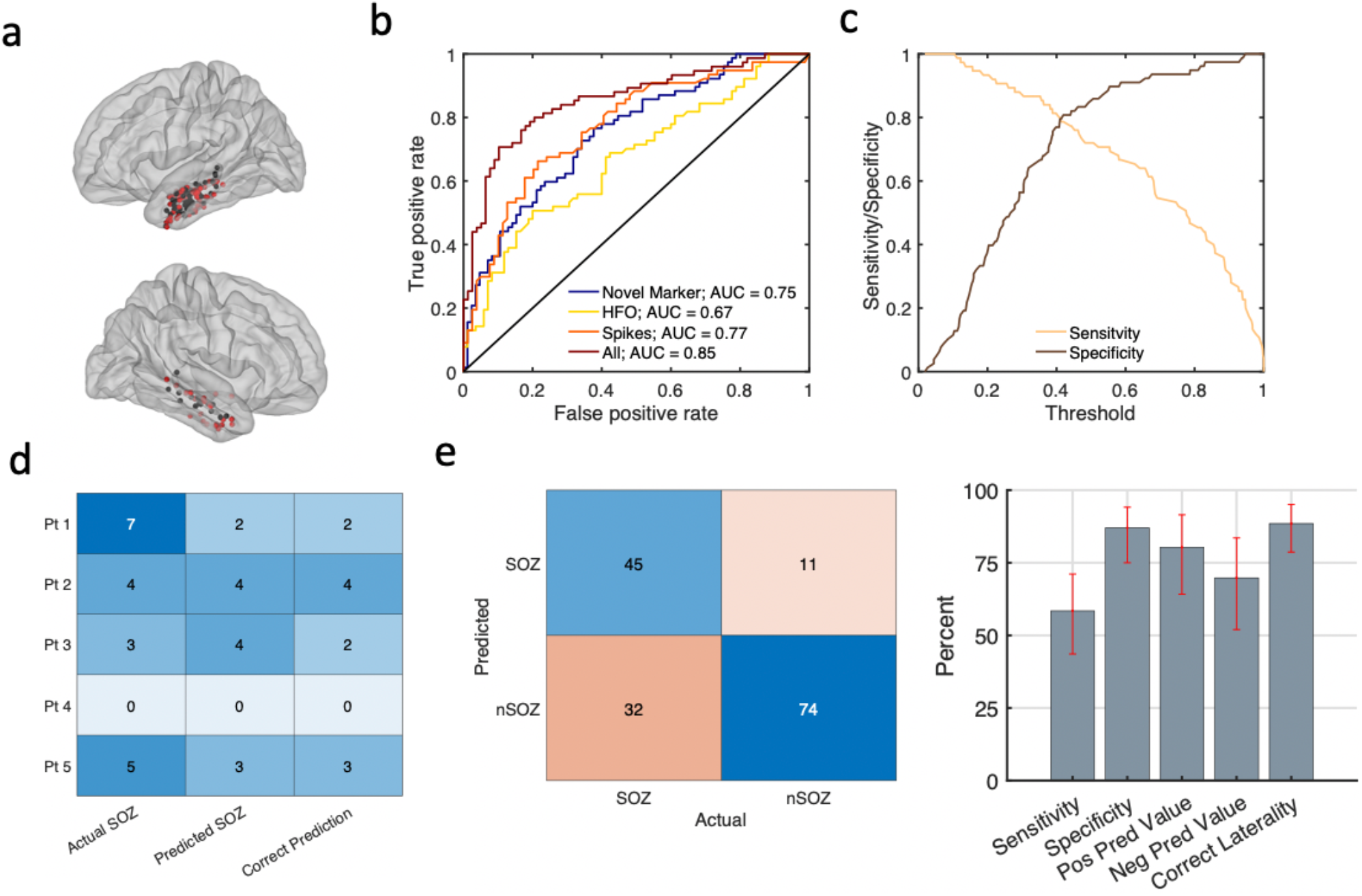
Classifying electrodes as SOZ or non-SOZ in the mesial temporal lobe. (a) 77 SOZ (red) and 85 non-SOZ (black) electrodes from 39 patients are displayed from the left (upper) and right (lower) mesial temporal brain regions. (b) Receiver Operating Characteristic (ROC) curve showing logistic regression classification for four models with features based on the novel marker, high frequency oscillations (HFOs), and interictal epileptiform spikes. Increased Area Under the Curve (AUC) reflects improved classification results (*p*<0.001 for AUC values). False positive rate equals one minus the specificity. True positive rate is equal to the sensitivity. (c) Sensitivity and specificity for varying threshold levels when all features are used for classification. (d) Predicted SOZ electrodes shown for example patients with threshold 0.6. Data for the tested patient is excluded from the training data, i.e. leave-one-patient-out cross-validation. (e) Predicted and Actual results for all 39 patients show sensitivity of 58%, specificity 87%, positive predictive value 80%, and negative predictive value 70%. For 89% of patients, lobar location of seizure onset zone (left, right, bilateral, or neither) was correctly predicted. Error bars are bootstrapped 95% confidence intervals.

To emphasize increased specificity, we choose a threshold of 0.6. Classifier results are shown for five individual example patients (Figure 4d). The classifier is not trained using data from the patient for whom results are shown, i.e. leave-one-patient-out cross validation was performed. Thus, these examples suggest real-world performance for a similar patient cohort. The specificity and positive predictive value of this classifier suggest high per patient accuracy. The classifier correctly lateralized the SOZ (i,e. left, right, bilateral, or neither) in 89% of patients using interictal data. Overall, specificity was 87% and positive predictive value was 80% for individual contacts.

We used a similar classifier to predict whether electrodes were involved in the SOZ for all neocortical electrodes (Figure 5). The high prevalence of non-SOZ electrodes compared to SOZ electrodes led to a high negative predictive value for this classifier with overall lower performance than for mesial temporal structures. For threshold 0.15 with leave-one-patient-out cross-validation, sensitivity was 41% and specificity was 75% with negative predictive value of 89%. In other words, although the classifier missed many SOZ electrodes, the prediction that an electrode was non-SOZ was 89% correct.

**Figure 5:**
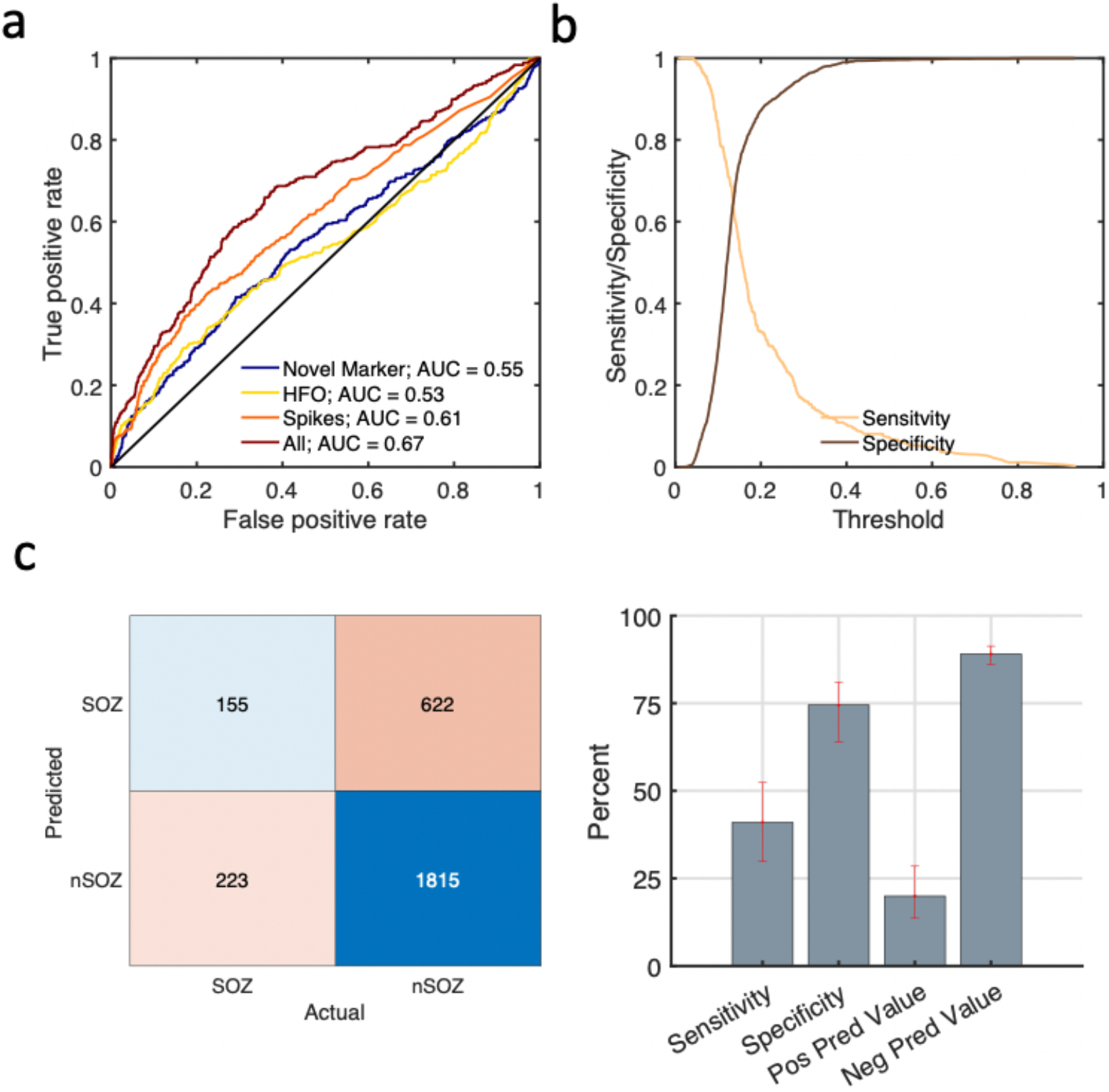
Classifying neocortical electrodes as SOZ or non-SOZ. (a) 378 SOZ and 2,437 non-SOZ electrodes were classified using logistic regression classification for four models with features based on the novel marker, high frequency oscillations (HFOs), and interictal epileptiform spikes. (b) Sensitivity and specificity for varying threshold levels when all features are used for classification. (c) Predicted and Actual results (threshold 0.15) show sensitivity of 41%, specificity 75%, positive predictive value 20%, and negative predictive value 89%. Error bars are bootstrapped 95% confidence intervals. Data for the tested patients is excluded from the training data, i.e. leave-one-patient-out cross-validation.

Given the overall promising classifier performance for SOZ electrodes, we wondered whether these interictal EEG markers could classify patients by outcome following surgical resection. Patients were divided into those with good outcomes (seizure-free or only auras, ILAE outcome scale 1 or 2) and poor outcomes (continued seizures, ILAE outcome scale 3 or greater). The addition of high frequency oscillations as a feature did not improve performance (sensitivity was the same and specificity was decreased to 87% from 91%). For each patient, the standard deviation of the Novel Marker values and the standard deviation of interictal spike rates across electrodes was used. Patients with good outcome had a greater variance of these interictal biomarkers across electrodes compared to poor outcome patients (Figure 6a). This may result from more optimal electrode coverage or more distinct spatial borders for good outcome patients that leads to greater differences between SOZ and non-SOZ brain regions. These two features are plotted against one another showing high specificity for good outcome patients (Figure 6b). Results from 10-fold cross-validation show that a predicted good outcome is rarely wrong (Figure 6c), with a positive predictive value of 92%. Thus, although good outcome patients were not always detected, if the model predicted a good outcome that prediction was likely correct.

**Figure 6:**
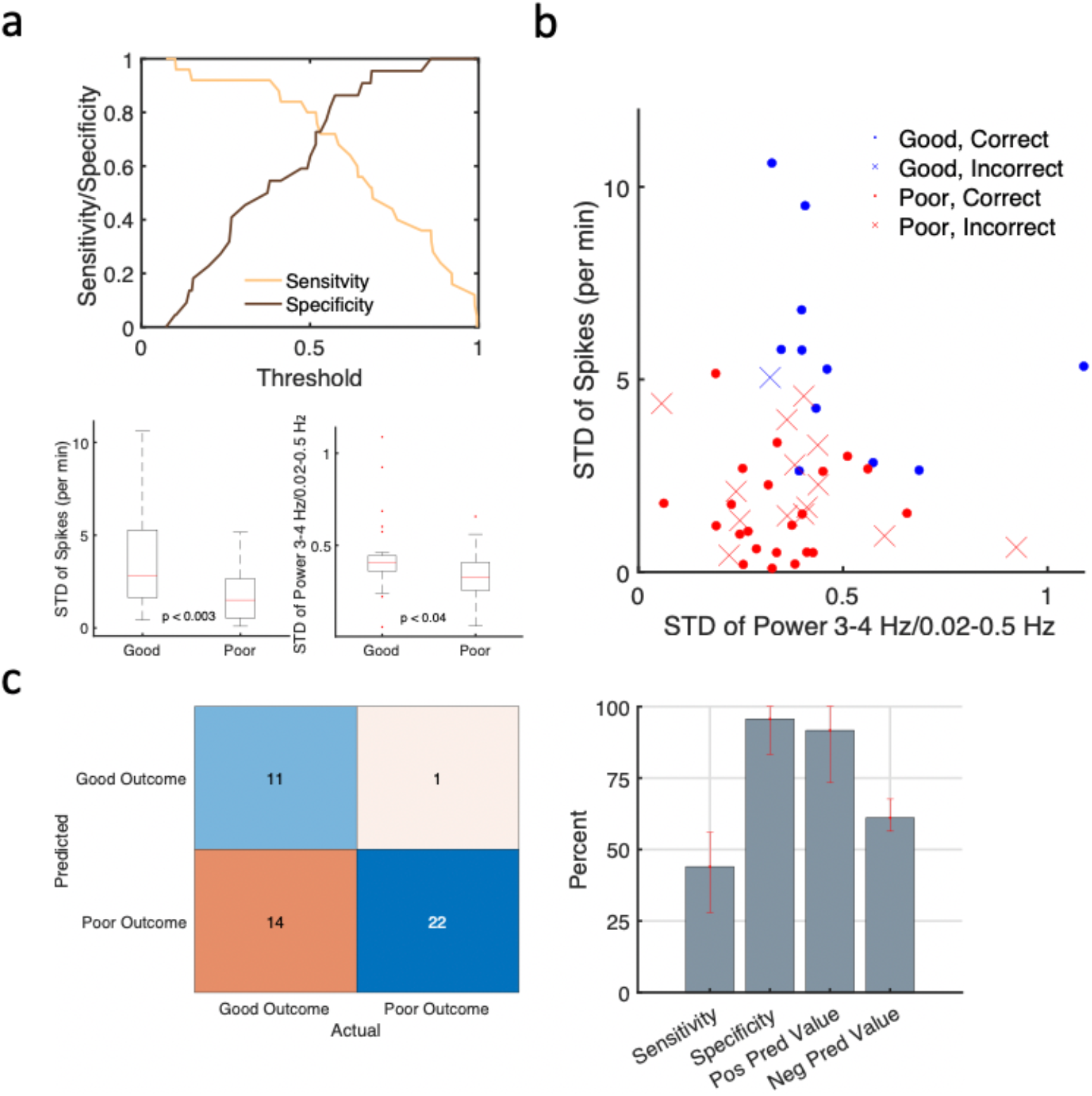
Classifying 48 patients as good (ILAE Outcome scale 1 or 2) or poor (ILAE >2) outcomes. (a) Sensitivity and specificity for varying threshold levels. Linear logistic regression classified patients using three features: the novel marker, spike amplitudes and reported baseline seizure frequency. The standard deviation of spikes and delta power across electrodes for a given patient was increased for patients with good vs poor outcomes. (b) Prediction (correct and incorrect) of patient outcome demonstrated for two features. (c) Predicted and Actual results (threshold 0.7) show sensitivity of 52%, specificity 91%, positive predictive value 87%, and negative predictive value 64%. Error bars are bootstrapped 95% confidence intervals. Results are for 10-fold cross-validation.

Finally, we wanted to gain further insight into the possible mechanisms underlying the decreased infraslow and increased fast activity for SOZ electrodes. We constructed a feed-forward neural network of 100 conductance-based neurons (Figure 7a). Each neuron included multiple timescale rate adaptation ^13^. Neuron outputs were filtered by a synapse with facilitation and two timescales of depression, consistent with physiologic data ^34^. The stimulus for this network was exponential-filtered Gaussian white noise with low and high standard deviations to simulate baseline and high firing rate conditions (Figure 7b and 7c). With the baseline stimulus, the simulated EEG power is similar to nSOZ electrodes (Figure 7d, black line). With the high firing rate stimulus, the overall power decreases, which may be counterintuitive and result from the filtering of synaptic depression (Figure 7d, blue line). Interestingly, infraslow activity is decreased while fast activity is increased relative to baseline when long-timescale adaptation is lost (Figure 7d, red line). Thus, the loss of long-timescale adaptation alters the power spectrum shape similar to results from Figures 2a and 3a, suggesting a possible relationship between infraslow activity and long-timescale adaptation.

**Figure 7:**
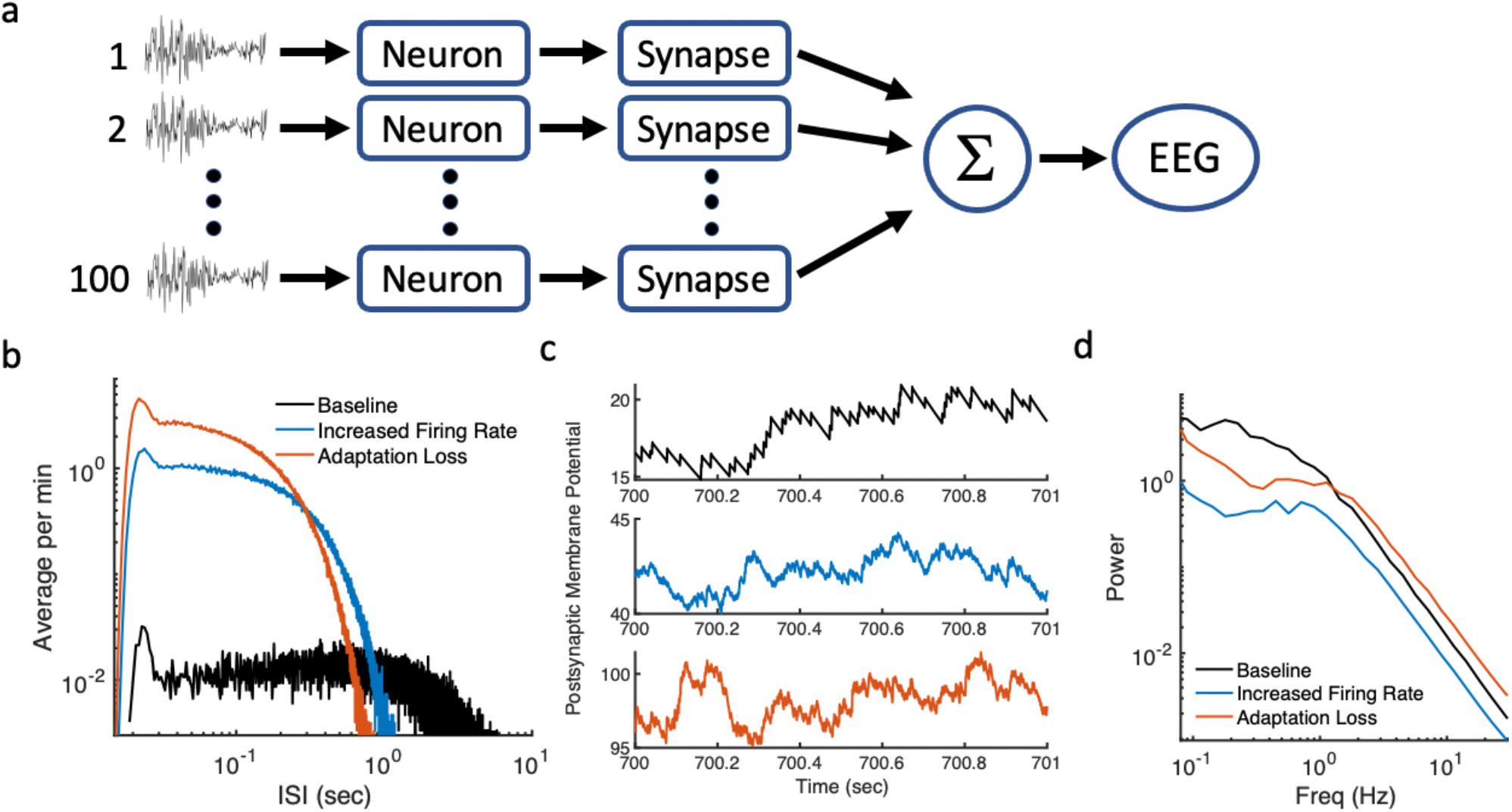
Neural network with loss of adaptation shows similar power spectra to SOZ electrodes. (a) Feed-forward neural network includes conductance-based neurons with multiple timescale adaptation and synapse with two timescales of synaptic depression. (b) Distributions of interspike intervals (ISIs). (c) Summed time-series following synaptic filtering. (d) Power spectra show increased high frequency power and decreased low frequency power compared to baseline.

## Discussion

We show that interictal EEG biomarkers including a novel marker comprised of a ratio of infraslow activity and faster EEG frequencies can predict seizure onset zones and patient outcomes. The novel marker is based on the observation that infraslow activity (0.02-0.5 Hz) is decreased near the SOZ and faster frequencies (2-50 Hz) are increased. A neural network with decreased long-timescale adaptation shows decreased infraslow activity. These findings support the hypothesis that distinct physiologic processes underlie infraslow activity, which may be related to inhibitory processes, while faster delta activity may be related to excitatory processes. In general, a power-based interictal marker such as the novel marker offers distinct advantages in terms of detection and computation. Combined with interictal spike rates and high frequency oscillation rates, these novel interictal features appear related to cortical excitability and offer predictive power related to seizure onset zone localization and patient outcomes following resection.

### Interictal Low Frequency Activity

There remains a need for interictal biomarkers of brain hyperexcitability. In clinical practice, the epileptogenic zone is approximated by recording multiple seizures to estimate the SOZ. However, recording a sufficient number of seizures to properly estimate the SOZ is often difficult and associated with increased time, expense, and patient morbidity. Interest in high frequency EEG phenomena such as HFOs has been driven in large part by the desire for a better interictal biomarker of hyperexcitability ^35^. Previous work suggests that increased rates of gamma range HFOs ^5,6^ and increased interictal epileptiform spike rates ^2,3,36^ are associated with seizure onset zones or hyperexcitable brain regions. Relatively less attention has been paid to lower frequency activity despite evidence that it modulates higher frequency activity ^1^, can be coupled to HFOs ^37^, and is related to neural mechanisms that modulate excitability ^14,16,17^.

We recently examined interictal data (recorded without DC-coupled amplifiers) from eight drug-resistant epilepsy patients with extensive subdural grid electrode coverage and found that 0.3-1 Hz activity was decreased near the seizure onset zone relative to higher frequency activity ^12^. This decreased activity was most evident during the sleep state but was also seen during the postictal state and wakefulness. 0.02-0.2 Hz activity modulates interictal epileptiform spikes ^1^. During sleep, an increase in interictal 1-4 Hz activity was seen in scalp EEG near the SOZ ^19^ and has been localized to the likely epileptogenic zone using source imaging ^38^.

### Underlying Mechanisms

Delta activity has been associated with increased excitability. During sleep increased delta activity appears to be a signature of hyperexcitable cortex, associated with seizures and interictal spikes ^19^.

According to the synaptic homeostatic hypothesis ^20^, increased neuronal activity requires increased synaptic renormalization, which then leads to increased delta activity during sleep. Delta activity has been associated with thalamocortical inputs, in contrast to activity less than 1 Hz that persisted without thalamic input ^17^.

Activity less than 1 Hz has been associated with modulation of cortical excitability, such as during sleep when there are alternating periods of excitation termed “up” and “down” states ^1,17^. One mechanism implicated in the determination of this slow oscillation frequency and the cessation of the “up” states ^39^ is related to activity-dependent potassium conductances; this single neuron mechanism regulates action potential firing via firing rate adaptation. A balance between the activity of these channels and incoming inhibition affected the slow oscillation frequency in brain slices ^16^. Firing rate adaptation and synaptic depression ^40,41^ are important for gain control, especially for activity with lower frequencies. These same mechanisms are modulated by many antiepileptic medications including those affecting sodium ^42^ or potassium channels ^40^. These results highlight the importance of firing rate adaptation and synaptic depression for controlling excitability.

Infraslow activity may be related to fractional differentiation, a form of multiple timescale feedback inhibition ^13–15^. Fractional differentiation results from a balanced form of inhibition in which a wide range of timescales are treated in a scale invariant manner. Thus, reduced infraslow activity may be associated with hyperexcitability.

Gamma oscillations have been associated with GABA-related networks, interneurons, inhibition, and pre-ictal activity ^43^. Interictal epileptiform spikes that were preceded by gamma oscillations were found to have a high association with the SOZ ^44^. Here, we measure power in the 20-50 Hz frequency range and not necessarily gamma oscillations. The increase in gamma power seen in the SOZ may reflect increased neural activity over a wide range of frequencies ^45^. HFOs have been postulated to reflect epileptogenic brain ^5^, although perhaps not on an individual patient level ^6^.

The reason for the relative decrease of infraslow activity in the SOZ electrodes remains unclear. Results from the neural network model suggest that decreased infraslow power is consistent with loss of long timescale action potential rate adaptation and synaptic depression in the setting of increased neural firing rates. Increased standard deviation of filtered white noise input has been taken to approximate the effects of increased input synchrony ^46^, as might be expected near the SOZ. When properly balanced, adaptation mechanisms yield scale invariant power law dynamics consistent with fractional differentiation ^13^ that can accentuate slow oscillations ^14^. Thus, loss of long-timescale adaptation is a reasonable explanation for decreased infraslow activity.

### Predicting Seizure Onset Zones

Localizing seizures remains challenging for patients with medication-resistant focal seizures. Patients are often admitted to the hospital for a week or more in order to record seizures for presurgical evaluation ^47^. Interictal biomarkers predicting, for example, laterality of involvement of mesial temporal lobes would be especially helpful. We showed that a combination of interictal biomarkers including the novel marker, spike rate, and HFO rate correctly predicted whether the left, right, both, or neither temporal lobes were involved for almost 90% of patients.

Quantification of HFOs has been the focus of a recently reported prospective trial where patient level predications were not helpful ^6^. Classification by HFO rates has yielded an Area Under the Curve (AUC) of 0.61 ^48^, while Support Vector Machine (SVM) classification using multiple HFO features from single channels yielded an AUC of 0.65. Here, the novel marker compares favorably with an AUC of 0.75 for mesial temporal lobe electrodes, compared to 0.67 for HFOs (Figure 4b). More sophisticated machine learning approaches with larger numbers of features can improve results ^7^. Here, we focused on a straightforward approach using logistic regression and a minimal number of features. In general, the Novel Marker seems to confer comparable but distinct diagnostic information as other markers. This physiology-based biomarker may be most useful in combination with other biomarkers, as has been suggested for HFOs and interictal spikes ^49^.

Ideally, one wants to know the epileptogenic zone rather than the seizure onset zone. Therefore, one may question the utility of predicting the seizure onset zone. Nonetheless, defining the seizure onset zone remains standard-of-care clinically. Thus, predicting the SOZ is of practical clinical benefit and, at least partially, overlaps with the epileptogenic zone. In terms of classification, the seizure onset zone is well-defined, in contrast to the epileptogenic zone. Even when accounting for patient outcomes in order to predict the epileptogenic zone, unknown details of resection size and resection boundaries in relation to the epileptogenic zone make it difficult to define as a concrete clinical tool.

### Predicting Patient Outcomes

One of the primary challenges in treating patients with irreversible therapeutic approaches such as surgical resection is determining on an individual patient level who may benefit. For this reason, additional information that may help determine whether benefits outweigh risks would improve patient care. The described interictal model has a high positive predictive value, which would indicate patients most likely to benefit from irreversible treatments. The ability of interictal markers to predict patient outcomes suggests that patient-level characteristics of the EEG differ between patient groups. Here, results suggest the variance of interictal biomarkers is increased when a single seizure onset zone is well-covered and bounded by non-SOZ electrodes.

### Limitations

Multiple factors may limit the predictive potential of low frequency activity. There is not necessarily a clear divide between different frequency bands. Multiple processes likely contribute to low frequency activity. Interictal epileptiform activity likely contributes to observed power changes, as the shape and frequency of spikes change near the SOZ. Interictal epileptiform activity contains a wide range of frequencies, including delta and beta activity, which affects multiple frequency bands. Although these data come from 1-3 am in early morning, sleep was not staged due to the absence of scalp data. Finally, neocortical data was recorded with subdural electrodes, whereas depth electrodes are now more commonly used; mesial temporal activity was recorded with depth electrodes.

### Conclusions

Interictal low frequency activity differs near hyperexcitable cortex and may reflect distinct underlying physiological processes related to cortical excitability. Continued work is needed to explore the utility of interictal biomarkers and low frequency activity for predicting cortical hyperexcitability and patient outcomes.

## Data Availability

De-identified data are available upon request.

## Acknowledgments

Hari Guragain and Tal Pal Attia for technical assistance and support. Mayo Clinic Division of Epilepsy for clinical support and care of patients. Christian Meisel, Tom Richner, and Larry Sorensen for helpful comments on draft manuscripts.

## References

1. Vanhatalo S, Palva JM, Holmes MD, et al. Infraslow oscillations modulate excitability and interictal epileptic activity in the human cortex during sleep [Internet]. Proc Natl Acad Sci U S A 2004;101(14):5053–5057.Available from: http://www.ncbi.nlm.nih.gov/pubmed/15044698

2. Staley KJ, White A, Dudek FE. Interictal spikes: harbingers or causes of epilepsy? [Internet]. Neurosci. Lett. 2011;497(3):247–50.[cited 2016 Oct 20] Available from: http://www.ncbi.nlm.nih.gov/pubmed/21458535

3. Lundstrom BN, Meisel C, Van Gompel J, et al. Comparing spiking and slow wave activity from invasive electroencephalography in patients with and without seizures [Internet]. Clin. Neurophysiol. 2018;129(5):909–919.[cited 2018 Jun 21] Available from: http://www.ncbi.nlm.nih.gov/pubmed/29550651

4. Marsh ED, Peltzer B, Brown III MW, et al. Interictal EEG spikes identify the region of electrographic seizure onset in some, but not all, pediatric epilepsy patients [Internet]. Epilepsia 2010;51(4):592–601.[cited 2020 Apr 10] Available from: http://www.ncbi.nlm.nih.gov/pubmed/19780794

5. Worrell GA, Parish L, Cranstoun SD, et al. High-frequency oscillations and seizure generation in neocortical epilepsy [Internet]. Brain 2004;127(7):1496–1506.[cited 2017 Apr 3] Available from: http://www.ncbi.nlm.nih.gov/pubmed/15155522

6. Jacobs J, Wu JY, Perucca P, et al. Removing high-frequency oscillations [Internet]. Neurology 2018;91(11):e1040–e1052.[cited 2018 Sep 12] Available from: http://www.ncbi.nlm.nih.gov/pubmed/30120133

7. Cimbalnik J, Klimes P, Sladky V, et al. Multi-feature localization of epileptic foci from interictal, intracranial EEG [Internet]. Clin. Neurophysiol. 2019;130(10):1945–1953.Available from: https://doi.org/10.1016/j.clinph.2019.07.024

8. Frauscher B, von Ellenrieder N, Zelmann R, et al. High-Frequency Oscillations in the Normal Human Brain [Internet]. Ann. Neurol. 2018;84(3):374–385.[cited 2020 May 29] Available from: http://doi.wiley.com/10.1002/ana.25304

9. Worrell GA, Gardner AB, Stead SM, et al. High-frequency oscillations in human temporal lobe: Simultaneous microwire and clinical macroelectrode recordings [Internet]. Brain 2008;131(4):928–937.[cited 2017 May 14] Available from: https://oup.silverchair-cdn.com/oup/backfile/Content_public/Journal/brain/131/4/10.1093/brain/awn006/2/awn006.pdf?Expires=1494891419&Signature=RJDVyvQss61BpXfV1D0ByxBo7wBNQbJQ7pZsqMHV8Tm5fnVZyKg2CZu7owRISUqUHo1UUxJq-Ra8kUZIeU~~TsPgWkHDzqvGLsaC1NAUXOINdgvg

10. Ikeda A, Taki W, Kunieda T, et al. Focal ictal direct current shifts in humanepilepsy as studied by subdural and scalp recording [Internet]. Brain 1999;122(5):827–838.[cited 2018 Sep 11] Available from: https://academic.oup.com/brain/article-lookup/doi/10.1093/brain/122.5.827

11. Constantino T, Rodin E. Peri-ictal and interictal, intracranial infraslow activity [Internet]. In: Journal of Clinical Neurophysiology. J Clin Neurophysiol; 2012 p. 298–308.[cited 2021 Feb 7] Available from: https://pubmed.ncbi.nlm.nih.gov/22854763/

12. Lundstrom BN, Boly M, Duckrow R, et al. Slowing less than 1 Hz is decreased near the seizure onset zone [Internet]. Sci. Rep. 2019;9(1):6218.[cited 2019 Apr 21] Available from: http://www.ncbi.nlm.nih.gov/pubmed/30996228

13. Lundstrom BN, Higgs MH, Spain WJ, Fairhall AL. Fractional differentiation by neocortical pyramidal neurons [Internet]. Nat Neurosci 2008;11(11):1335–1342.Available from: http://www.ncbi.nlm.nih.gov/entrez/query.fcgi?cmd=Retrieve&db=PubMed&dopt=Citation&list_uids=18931665

14. Lundstrom BN. Modeling multiple time scale firing rate adaptation in a neural network of local field potentials [Internet]. J. Comput. Neurosci. 2015;38(1):189–202.Available from: http://link.springer.com/10.1007/s10827-014-0536-2

15. Lundstrom BN, Fairhall AL, Maravall M. Multiple timescale encoding of slowly varying whisker stimulus envelope in cortical and thalamic neurons in vivo [Internet]. J Neurosci 2010;30(14):5071–5077.Available from: http://www.ncbi.nlm.nih.gov/pubmed/20371827

16. Sanchez-Vives M V., Mattia M, Compte A, et al. Inhibitory Modulation of Cortical Up States [Internet]. J. Neurophysiol. 2010;104(3):1314–1324.[cited 2018 Aug 22] Available from: http://www.physiology.org/doi/10.1152/jn.00178.2010

17. Steriade M. Grouping of brain rhythms in corticothalamic systems [Internet]. Neuroscience 2006;137(4):1087–1106.Available from: http://www.ncbi.nlm.nih.gov/pubmed/16343791

18. Podvalny E, Noy N, Harel M, et al. A unifying principle underlying the extracellular field potential spectral responses in the human cortex. [Internet]. J. Neurophysiol. 2015;114(1):505–19.[cited 2018 Oct 29] Available from: http://www.ncbi.nlm.nih.gov/pubmed/25855698

19. Boly M, Jones B, Findlay G, et al. Altered sleep homeostasis correlates with cognitive impairment in patients with focal epilepsy [Internet]. Brain 2017;140(4):1026–1040.[cited 2017 May 8] Available from: http://www.ncbi.nlm.nih.gov/pubmed/28334879

20. Tononi G, Cirelli C. Sleep and the Price of Plasticity: From Synaptic and Cellular Homeostasis to Memory Consolidation and Integration [Internet]. Neuron 2014;81(1):12–34.[cited 2018 Jun 21] Available from: http://www.ncbi.nlm.nih.gov/pubmed/24411729

21. Ebersole J, Husain A, Nordli D. Current Practice of Clinical Electroencephalography. Fourth. Wolters Kluwer; 2014.

22. Engel J, McDermott MP, Wiebe S, et al. Early surgical therapy for drug-resistant temporal lobe epilepsy: A randomized trial [Internet]. JAMA - J. Am. Med. Assoc. 2012;307(9):922–930.[cited 2021 Jan 17] Available from: https://jamanetwork.com/

23. King-Stephens D, Mirro E, Weber PB, et al. Lateralization of mesial temporal lobe epilepsy with chronic ambulatory electrocorticography [Internet]. Epilepsia 2015;56(6):959–967.[cited 2019 Jan 14] Available from: http://www.ncbi.nlm.nih.gov/pubmed/25988840

24. Hirsch LJ, Mirro EA, Salanova V, et al. Mesial temporal resection following long-term ambulatory intracranial EEG monitoring with a direct brain-responsive neurostimulation system [Internet]. Epilepsia 2020;61(3):408–420.[cited 2021 Jan 17] Available from: https://pubmed.ncbi.nlm.nih.gov/32072621/

25. Guragain H, Cimbalnik J, Stead M, et al. Spatial variation in high-frequency oscillation rates and amplitudes in intracranial EEG. Neurology 2018;90(8):E639–E646.

26. Papademetris X, Jackowski MP, Rajeevan N, et al. BioImage Suite: An integrated medical image analysis suite: An update. [Internet]. Insight J. 2006;2006:209.[cited 2020 Apr 2] Available from: http://www.ncbi.nlm.nih.gov/pubmed/25364771

27. Groppe DM, Bickel S, Dykstra AR, et al. iELVis: An open source MATLAB toolbox for localizing and visualizing human intracranial electrode data [Internet]. J. Neurosci. Methods 2017;281:40–48.[cited 2020 Apr 2] Available from: https://www.sciencedirect.com/science/article/pii/S0165027017300365

28. Desikan RS, Ségonne F, Fischl B, et al. An automated labeling system for subdividing the human cerebral cortex on MRI scans into gyral based regions of interest [Internet]. Neuroimage 2006;31(3):968–980.[cited 2020 Apr 2] Available from: http://www.ncbi.nlm.nih.gov/pubmed/16530430

29. Cimbálník J, Hewitt A, Worrell G, Stead M. The CS algorithm: A novel method for high frequency oscillation detection in EEG [Internet]. J. Neurosci. Methods 2018;293:6–16.[cited 2020 Apr 2] Available from: http://www.ncbi.nlm.nih.gov/pubmed/28860077

30. Mitra PP, Bokil H. Observed Brain Dynamics. Oxford Univ Press; 2007.

31. Barkmeier DT, Shah AK, Flanagan D, et al. High inter-reviewer variability of spike detection on intracranial EEG addressed by an automated multi-channel algorithm. [Internet]. Clin. Neurophysiol. 2012;123(6):1088–95.[cited 2016 Oct 20] Available from: http://www.ncbi.nlm.nih.gov/pubmed/22033028

32. James G, Witten D, Hastie T, Tibshirani R. An Introduction to Statistical Learning with Applications in R. Springer; 2013.

33. Press WH. Numerical recipes in C : the art of scientific computing [Internet]. 2nd ed. Cambridge ; New York: Cambridge University Press; 1992.Available from: http://www.loc.gov/catdir/description/cam026/94105607.html

34. Varela JA, Sen K, Gibson J, et al. A quantitative description of short-term plasticity at excitatory synapses in layer 2/3 of rat primary visual cortex [Internet]. J Neurosci 1997;17(20):7926–7940.Available from: http://www.ncbi.nlm.nih.gov/entrez/query.fcgi?cmd=Retrieve&db=PubMed&dopt=Citation&list_uids=9315911

35. Jacobs J, Staba R, Asano E, et al. High-frequency oscillations (HFOs) in clinical epilepsy. [Internet]. Prog. Neurobiol. 2012;98(3):302–15.[cited 2017 Apr 10] Available from: http://linkinghub.elsevier.com/retrieve/pii/S0301008212000330

36. Stead M, Bower M, Brinkmann BH, et al. Microseizures and the spatiotemporal scales of human partial epilepsy. [Internet]. Brain 2010;133(9):2789–97.[cited 2016 Oct 20] Available from: http://www.ncbi.nlm.nih.gov/pubmed/20685804

37. Amiri M, Frauscher B, Gotman J. Interictal coupling of HFOs and slow oscillations predicts the seizure-onset pattern in mesiotemporal lobe epilepsy [Internet]. Epilepsia 2019;60(6):1160–1170.[cited 2021 Apr 28] Available from: https://pubmed.ncbi.nlm.nih.gov/31087662/

38. Baldini S, Coito A, Korff CM, et al. Localizing non-epileptiform abnormal brain function in children using high density EEG: Electric Source Imaging of focal slowing. Epilepsy Res. 2020;159(November 2019)

39. Neske GT. The Slow Oscillation in Cortical and Thalamic Networks: Mechanisms and Functions. [Internet]. Front. Neural Circuits 2015;9:88.[cited 2017 Aug 1] Available from: http://www.ncbi.nlm.nih.gov/pubmed/26834569

40. Benda J, Herz A V. A universal model for spike-frequency adaptation [Internet]. Neural Comput 2003;15(11):2523–2564.Available from: http://www.ncbi.nlm.nih.gov/entrez/query.fcgi?cmd=Retrieve&db=PubMed&dopt=Citation&list_uids=14577853

41. Puccini GD, Sanchez-Vives M V, Compte A. Integrated mechanisms of anticipation and rate-of-change computations in cortical circuits [Internet]. PLoS Comput Biol 2007;3(5):e82.Available from: http://www.ncbi.nlm.nih.gov/entrez/query.fcgi?cmd=Retrieve&db=PubMed&dopt=Citation&list_uids=17500584

42. de Biase S, Gigli GL, Valente M, Merlino G. Lacosamide for the treatment of epilepsy [Internet]. Expert Opin Drug Metab Toxicol 2014;10(3):459–468.Available from: http://www.ncbi.nlm.nih.gov/pubmed/24479852

43. Hughes JR. Gamma, fast, and ultrafast waves of the brain: Their relationships with epilepsy and behavior. Epilepsy Behav. 2008;13(1):25–31.

44. Ren L, Kucewicz MT, Cimbalnik J, et al. Gamma oscillations precede interictal epileptiform spikes in the seizure onset zone. [Internet]. Neurology 2015;84(6):602–8.[cited 2015 Oct 27] Available from: http://www.ncbi.nlm.nih.gov/pubmed/25589669

45. Miller KJ, Sorensen LB, Ojemann JG, Den Nijs M. Power-law scaling in the brain surface electric potential. PLoS Comput. Biol. 2009;5(12)

46. Moreno R, de la Rocha J, Renart A, Parga N. Response of spiking neurons to correlated inputs [Internet]. Phys Rev Lett 2002;89(28 Pt 1):288101.Available from: http://www.ncbi.nlm.nih.gov/entrez/query.fcgi?cmd=Retrieve&db=PubMed&dopt=Citation&list_uids=12513181

47. Gazzola DM, Thawani S, Agbe-Davies O, Carlson C. Epilepsy monitoring unit length of stay [Internet]. Epilepsy Behav. 2016;58:102–105.[cited 2021 Jan 19] Available from: https://pubmed.ncbi.nlm.nih.gov/27064830/

48. Cimbalnik J, Brinkmann B, Kremen V, et al. Physiological and pathological high frequency oscillations in focal epilepsy. Ann. Clin. Transl. Neurol. 2018;5(9):1062–1076.

49. Roehri N, Pizzo F, Lagarde S, et al. High-Frequency Oscillations Are Not Better Biomarkers of Epileptogenic Tissues Than Spikes. Ann. Neurol. 2018;(83):84–97.

